# Protocol for a nationwide Internet-based health survey in workers during the COVID-19 pandemic in 2020

**DOI:** 10.1101/2021.02.02.21249309

**Authors:** Yoshihisa Fujino, Tomohiro Ishimaru, Hisashi Eguchi, Mayumi Tsuji, Seiichiro Tateishi, Akira Ogami, Koji Mori, Shinya Matsuda, for the CORoNaWork project

## Abstract

The ever-changing social implications of the COVID-19 pandemic have resulted in an urgent need to understand the working environments and health status of workers. We conducted a nationwide Internet-based health survey in Japanese workers in December 2020, in the midst the country’s “third wave” of COVID-19 infection. Of 33,087 surveys collected, 6,051 were determined to have invalid responses. The 27,036 surveys included in the study were balanced in terms of geographical area, participant sex, and type of work, according to the sampling plan. Men were more likely than women to have telecommuted, while women were more likely to have resigned since April 2020. Moreover, 40% and 9.1% of respondents had a K6 score of 5 or higher and 13 or higher, respectively, they did not exhibit extremely poor health. The present study describes the protocol used to conduct an Internet-based health survey in workers and a summary of its results during a period when COVID-19 was spreading rapidly in Japan. In the future, we plan to use this survey to examine the impact of COVID-19 on workers’ work styles and health.

## Introduction

The global outbreak of COVID-19 in 2020 has had an enormous impact on the economy, daily life, and medical practice in Japan[1–3]. In April 2020, the Japanese government declared a state of emergency and asked the population to refrain from going out and for workplaces to close. These broad restrictions on movement, which aimed to control the pandemic, reduced economic activity, which in turn caused a deterioration in work environment, worsening of corporate financial performance, and increases in layoffs and unemployment[4]. Between February and December 2020, the COVID-19 pandemic caused more than 800 companies to declare bankruptcy[5].

The COVID-19 pandemic has brought about dramatic changes to the work environment. One major change is the wide adoption of telecommuting, which was boosted by the government’s state of emergency declaration in April[6,7]. In Japan, telecommuting was previously discussed as a strategy for reducing long working hours[8]. The COVID-19 pandemic and the state of emergency declaration pushed many companies in Japan to rapidly adopt telecommuting[6,7]. While the health effects of telecommuting on workers have not been fully clarified, many experts have expressed concern about the impact on lifestyle habits such as alcohol consumption, exercise habits, and dietary habits. There are also concerns about the impact on musculoskeletal diseases, back pain, and video display terminal-related diseases in the home environment, which are inadequately managed compared to those occurring in the office environment.

In December 2020, Japan experienced its “third wave” of infections, the largest increase to date even compared to the previous waves experienced in May and August. On December 22, the number of infections in the country reached a record high of approximately 3,200. On December 15, the Ministry of Health, Labor and Welfare announced that seven prefectures had reached Stage 4 of the government’s four-stage alert scale, indicating that occupancy of hospital beds reserved for the severely ill had exceeded 50%, and that the medical supply system was reaching its limit. On December 21, the Japan Medical Association declared a medical emergency. In a statement, it announced that patients infected with COVID-19 and regular people in Japan would not be able to receive normal medical care, and that all necessary medical care provisions across the country would be brought to a standstill[9].

The ever-changing landscape and impact of the COVID-19 pandemic has resulted in an urgent need to understand the working and social environment and health status of workers. A number of concerns are emerging, including those related to the socioeconomic status, mental health, lifestyle, work productivity, isolation and loneliness, family relationships, infection anxiety, and infection prevention activities of workers, and to the corporate support systems and corporate infection prevention measures put in place during the COVID-19 pandemic. We examined some of these by conducting an urgent large-scale Internet survey of workers in the midst of the third wave of COVID-19 infection in Japan in December 2020.

## Methods

This survey is a prospective cohort study conducted online among Internet monitors. The baseline survey was conducted from December 22 to 25, 2020. A second survey is scheduled for 2021. The study targeted those who were working and between the ages of 20 and 65 at the time of the baseline survey, and was approved by the Ethics Committee of the University of Occupational and Environmental Health, Japan.

### Sampling plan

To avoid geographic bias among participants, it was necessary to adopt a sampling plan that accounted for regional characteristics. However, because some prefectures only had a few registered monitors, sampling by prefecture was not possible. Therefore, the prefectures were divided into five regions based on geographic region and infection status: the prefectures were divided into four regions based on the cumulative infection rate, and the region with the highest cumulative infection rate was further divided into the Kanto region and non-Kanto region (Table 1). The cumulative infection rate was based on information available as of December 16, 2020.

**Table 1.** Surveys collected based on sampling plan

| Region/Prefecture | Cumulative COVID-19<br>incidence rate per<br>million population | Total<br>(n=33,087) | Office workers |  | Non-office workers |  |
| --- | --- | --- | --- | --- | --- | --- |
|  |  |  | Male<br>(n=8,261) | Female<br>(n=8,300) | Male<br>(n=8,323) | Female<br>(n=8,203) |
| (Kanto region) Tokyo*, Kanagawa, Saitama, Chiba | 1168-3496 | 6,657 | 1,682 | 1,684 | 1,651 | 1,640 |
| (non-Kanto region) Okinawa, Osaka*, Hokkaido*, Aichi*,<br>Hyogo*, Fukuoka, Kyoto, Nara |  | 6,700 | 1,654 | 1,696 | 1,676 | 1,674 |
| Gunma, Ishikawa, Gifu, Kumamoto, Ibaragi, Miyagi,<br>Hiroshima, Shiga, Mie*, Kochi*, Sizuoka, Wakayama,<br>Miyazaki, Yamanashi, Kagoshima | 535-911 | 6,579 | 1,652 | 1,639 | 1,659 | 1,629 |
| Nagano, Saga, Tochigi, Oita, Toyama, Okayama, Fukui | 438-490 | 6,537 | 1,627 | 1,620 | 1,665 | 1,625 |
| Fukushima, Yamaguchi, Aomori, Ehime, Yamagata,<br>Nagasaki, Iwate, Tokushima, Shimane, Kagawa, Nigata,<br>Tottori, Akita | 97-356 | 6,614 | 1,646 | 1,661 | 1,672 | 1,635 |

\* Prefectures that had reached Stage 4 of the government's four-stage alert scale, indicating that occupancy of hospital beds reserved for the severely ill had exceeded 50%, according to the Ministry of Health, Labor and Welfare on December 15, 2020.

The sampling plan was designed to collect an equal number of respondents from across 20 collection units, each consisting of a combination of five regions, with comparable sex, and office and non-office worker status. The target sample size was 30,000, with 1,500 respondents from each collection unit. A total of 1,650 respondents, which represents the target sample size plus a margin of 10%, were collected from each collection unit. Ultimately, a total of 33,087 respondents were collected for the Internet survey.

### Subject recruitment procedure

The survey was commissioned by Cross Marketing Inc. (Tokyo, Japan), which has 4.7 million registered monitors. Of the registered monitors, 605,381 were sent an invitation to participate via e-mail. Of these, a total of 55,045 registered monitors answered the initial screening questions to participate in the survey, and 33,302 who matched the survey’s criteria (worker status, region, sex, and age) responded to the survey.

The survey was launched on December 22, 2020, and by December 26, 33,302 people had participated. Approximately 98% of the sample was collected by December 23. Collection of the remaining sample, which consisted of women only, was completed on December 26.

### Data retrieval

Initially, of the 33,302 respondents, 215 were excluded because they were deemed to have provided fraudulent responses by Cross Marketing Inc., leaving 33,087 respondents. Subsequently, 6,051 surveys determined to contain invalid responses or response errors were excluded, leaving 27,036 samples for inclusion in the study. The exclusion criteria were as follows: extremely short response time (≤6 minutes), extremely low body weight (<30 kg), extremely short height (<140 cm), inconsistent answers to similar questions throughout the survey (e.g., inconsistency to questions about marital status and living area), and wrong answers to a staged question used to identify fraudulent responses (choose the third largest number from the following five numbers).

### Measurements

The survey items included basic socio-demographic characteristics such as family structure, income, educational background, area of residence, area of employment, and work environment-related factors. The survey included work-related questionnaires like the Japanese version of the Job Content Questionnaire[10,11], the Japanese version of the 3-item Utrecht Work Engagement Scale [12,13], and Work Functioning Impairment Scale (WFun)[14], and inquired about frequency of working at home. Psychosocial conditions were examined through assessment of health-related quality of life (HRQOL), K6[15], and loneliness. HRQOL was measured using the CDC HRQOL-4[16,17], which was originally developed by the US Centers for Disease Control and Prevention. Health-related items included medical history, treatment interruptions, back pain, and stiff shoulders. Lifestyle-related items included items related to smoking, drinking, exercise, and eating habits. The survey also asked about preventive behaviors against infection, such as hand washing and gargling, and concerns about infection.

## Results

Target sample sizes were successfully obtained for all allocation conditions, including with regard to region, sex, and type of work (Table 1).

Table 2 shows the number of subjects included for further analyses and the number of surveys judged to contain fraudulent responses, by sampling unit and sex. There was no significant regional difference in the percentage of responses that were judged to be fraudulent.

**Table 2.** Number of survey responses eligible for analysis and the number judged to have invalid responses.

| Region/Prefecture | Samples for analysis |  | Samples judged to have invalid responses |  |  |  |
| --- | --- | --- | --- | --- | --- | --- |
|  | Male<br>(n=13,814) | Female<br>(n=13,222) | Male<br>(n=2,770) | Female<br>(n=3,281) | % |  |
|  |  |  |  |  | Male | Female |
| (Kanto region) Tokyo*, Kanagawa, Saitama, Chiba | 2,831 | 2,629 | 502 | 695 | 18% | 26% |
| (non-Kanto region) Okinawa, Osaka*, Hokkaido*,<br>Aichi*, Hyogo*, Fukuoka, Kyoto, Nara | 2,783 | 2,667 | 547 | 703 | 20% | 26% |
| Gunma, Ishikawa, Gifu, Kumamoto, Ibaragi, Miyagi,<br>Hiroshima, Shiga, Mie*, Kochi*, Sizuoka, Wakayama,<br>Miyazaki, Yamanashi, Kagoshima | 2,725 | 2,609 | 586 | 659 | 22% | 25% |
| Nagano, Saga, Tochigi, Oita, Toyama, Okayama, Fukui | 2,766 | 2,684 | 526 | 561 | 19% | 21% |
| Fukushima, Yamaguchi, Aomori, Ehime, Yamagata,<br>Nagasaki, Iwate, Tokushima, Shimane, Kagawa, Nigata,<br>Tottori, Akita | 2,709 | 2,633 | 609 | 663 | 22% | 25% |
| Subtotal | 13814 | 13222 | 2770 | 3281 | 20% | 25% |

Table 3 compares the characteristics of respondents who were included and excluded from the analysis. The following question was used to detect fraudulent responses: “Choose the third largest number from the following five numbers.” We compared the characteristics of those who answered this question correctly versus incorrectly. Of those who answered incorrectly, 1.2% had extremely low body weight and 0.7% had extremely short height; both of these were significantly more prevalent than among respondents who answered correctly. Those who answered incorrectly were also more likely to provide inconsistent answers related to cohabitants and residence, and to have extremely short response times, compared to those who answered correctly. Conversely, people with extremely short response times were more likely than those with appropriate response times to answer the fraud-detecting question incorrectly, or to give inconsistent answers to questions about cohabitants and residence, or to have extremely low body weight or extremely short height.

**Table 3.** Comparison of analyzed and excluded samples

|  | Samples for analysis |  |  | Response to question aimed at detecting fraudulent responses |  |  | Time taken to respond |  |  |
| --- | --- | --- | --- | --- | --- | --- | --- | --- | --- |
|  | Samples for analysis<br>n=27036 | Samples judged to have<br>invalid responses<br>n=6051 | *p* | Correct<br>n=30652 | Incorrect<br>n=2435 | *p* | >6 min<br>n=30688 | ≤6 min<br>n=2399 | *p* |
| Age, mean (SD) | 47.0 (10.5) | 42.8 (10.9) | <0.001 | 46.5 (10.6) | 42.9 (11.4) | <0.001 | 46.7 (10.6%) | 40.4 (10.0%) | <0.001 |
| Sex, male (%) | 13814 (51.1%) | 2770 (45.8%) | <0.001 | 15631 (51.0%) | 953 (39.1%) | <0.001 | 15381 (50.1%) | 1203 (50.1%) | 0.980 |
| Weight <30kg (%) | 0 (0.0%) | 101 (1.7%) | <0.001 | 72 (0.2%) | 29 (1.2%) | <0.001 | 77 (0.3%) | 24 (1.0%) | <0.001 |
| Height <140cm (%) | 0 (0.0%) | 71 (1.2%) | <0.001 | 55 (0.2%) | 16 (0.7%) | <0.001 | 58 (0.2%) | 13 (0.5%) | <0.001 |
| Incorrect answer to question aimed<br>at detecting fraudulent responses (%) | 0 (0.0%) | 2435 (40.2%) | <0.001 | 0 (0.0%) | 2435 (100.0%) | <0.001 | 2080 (6.8%) | 355 (14.8%) | <0.001 |
| Inconsistent responses regarding<br>family members living together (%) | 0 (0.0%) | 184 (3.0%) | <0.001 | 145 (0.5%) | 39 (1.6%) | <0.001 | 138 (0.4%) | 46 (1.9%) | <0.001 |
| Inconsistent responses regarding<br>area of residence (%) | 0 (0.0%) | 1852 (30.6%) | <0.001 | 1592 (5.2%) | 260 (10.7%) | <0.001 | 1525 (5.0%) | 327 (13.6%) | <0.001 |
| Time taken to respond ≤6 minutes<br>(%) | 0 (0.0%) | 2399 (39.6%) | <0.001 | 2044 (6.7%) | 355 (14.6%) | <0.001 | 0 (0.0%) | 2399 (100.0%) | <0.001 |

Table 4 shows the characteristics of the analysis subjects by sampling unit. Region 5, corresponding to the Kanto region, which had the highest cumulative infection rate, had more high-income earners and more people with telecommuting experience than the other regions. Region 5 also had more people with high WFun scores, high K6 scores, and poor self-rated health. In addition, 54 (1%) respondents from Region 5 reported a history of COVID-19 infection, compared to 30 (0.6%) respondents from Region 1.

**Table 4.** Basic characteristics of respondents by region

|  | Sampling unit (n=27036) |  |  |  |  | p-value |
| --- | --- | --- | --- | --- | --- | --- |
|  | Region 1 | Region 2 | Region 3 | Region 4 | Region 5 |  |
| N | 5342 | 5450 | 5334 | 5450 | 5460 |  |
| Age, mean | 46.5 (10.7) | 45.8 (10.8) | 47.1 (10.5) | 47.8 (10.3) | 47.7 (10.3) | <0.001 |
| Sex, male (%) | 2709 (50.7%) | 2766 (50.8%) | 2725 (51.1%) | 2783 (51.1%) | 2831 (51.8%) | 0.770 |
| Marriage status |  |  |  |  |  |  |
| Currently married | 3022 (56.6%) | 3211 (58.9%) | 2999 (56.2%) | 2938 (53.9%) | 2859 (52.4%) | <0.001 |
| Divorced or widowed | 586 (11.0%) | 588 (10.8%) | 575 (10.8%) | 601 (11.0%) | 493 (9.0%) |  |
| Never married | 1734 (32.5%) | 1651 (30.3%) | 1760 (33.0%) | 1911 (35.1%) | 2108 (38.6%) |  |
| Household income |  |  |  |  |  |  |
| Less than 2 million yen | 385 (7.2%) | 271 (5.0%) | 368 (6.9%) | 378 (6.9%) | 307 (5.6%) | <0.001 |
| 2 to 9.99 million yen | 4301 (80.5%) | 4409 (80.9%) | 4293 (80.5%) | 4409 (80.9%) | 4419 (80.9%) |  |
| More than 10 million yen | 656 (12.3%) | 795 (14.6%) | 767 (14.4%) | 843 (15.5%) | 1189 (21.8%) |  |
| Job type |  |  |  |  |  |  |
| Mainly desk work (clerical or computer work) | 2689 (50.3%) | 2684 (49.2%) | 2626 (49.2%) | 2701 (49.6%) | 2768 (50.7%) | <0.001 |
| Mainly talking to people (customer service, sales, selling, etc.) | 1287 (24.1%) | 1315 (24.1%) | 1304 (24.4%) | 1474 (27.0%) | 1547 (28.3%) |  |
| Mainly labor (work at production sites, physical work, nursing care, etc.) | 1366 (25.6%) | 1451 (26.6%) | 1404 (26.3%) | 1275 (23.4%) | 1145 (21.0%) |  |
| Current smoker, % | 1410 (26.4%) | 1302 (23.9%) | 1386 (26.0%) | 1418 (26.0%) | 1488 (27.3%) | 0.002 |
| Do you telecommute? (Almost never) | 4641 (86.9%) | 4632 (85.0%) | 4382 (82.2%) | 4174 (76.6%) | 3447 (63.1%) |  |
| Have you resigned or changed jobs since April 2020? (Yes) | 320 (6.0%) | 326 (6.0%) | 369 (6.9%) | 375 (6.9%) | 331 (6.9%) | 0.360 |
| Do you need any consideration or support from your company to continue working in your current health condition? (Yes) | 1369 (25.6%) | 1395 (25.6%) | 1353 (25.4%) | 1339 (24.6%) | 1319 (24.2%) | 0.600 |
| Have you been infected with COVID-19? (Yes) | 30 (0.6%) | 27 (0.5%) | 38 (0.7%) | 46 (0.8%) | 54 (1.0%) | 0.015 |
| WFun≥21 | 1208 (22.6%) | 1193 (21.9%) | 1189 (22.3%) | 1098 (20.1%) | 1103 (20.2%) | 0.001 |
| K6≥5 | 2195 (41.1%) | 2256 (41.4%) | 2180 (40.9%) | 2064 (37.9%) | 2122 (38.9%) | 0.004 |
| K6≥10 | 1050 (19.7%) | 992 (18.2%) | 1033 (19.4%) | 990 (18.2%) | 984 (18.0%) | 0.080 |
| K6≥13 | 519 (9.7%) | 470 (8.6%) | 551 (10.3%) | 455 (8.3%) | 465 (8.5%) | <0.001 |
| Perceived poor self-rated health | 2811 (52.6%) | 2781 (51.0%) | 2723 (51.0%) | 2639 (48.4%) | 2626 (48.1%) | <0.001 |

Table 5 summarizes the characteristics of the analysis subjects by sex. The sample size was balanced for sex and type of work according to the study design. Men accounted for 51% of the total sample. Office workers accounted for 49%, among both men and women. The smoking rate among men was 35.1%, higher than that among women (16.3%). Men were more likely than women to have telecommuted, while women were more likely to have resigned since April 2020. A total of 0.7% of both men and women reported a history of COVID-19 infection.

**Table 5.** Basic characteristics of respondents by sex

|  | Total | Sex |  |
| --- | --- | --- | --- |
|  |  | Male | Female |
| N | 27036 | 13814 | 13222 |
| Age, mean | 47.0 (10.5) | 51.52 (8.5) | 42.3 (10.4) |
| Sex, male (%) | 13814 (51.1%) | 13814 (100.0%) | - |
| Marriage status |  |  |  |
| Currently married | 15029 (55.6%) | 9449 (68.4%) | 5580 (42.2%) |
| Divorced or widowed | 2843 (10.5%) | 981 (7.1%) | 1862 (14.1%) |
| Never married | 9164 (33.9%) | 3384 (24.5%) | 5780 (43.7%) |
| Household income |  |  |  |
| Less than 2 million yen | 1709 (6.3%) | 705 (5.1%) | 1004 (7.6%) |
| 2 to 9.99 million yen | 21077 (78.0%) | 10561 (76.5%) | 10516 (79.5%) |
| More than 10 million yen | 4250 (15.7%) | 2548 (18.4%) | 1702 (12.9%) |
| Job type |  |  |  |
| Mainly desk work (clerical or computer work) | 13468 (49.8%) | 6896 (49.9%) | 6572 (49.7%) |
| Mainly talking to people (customer service, sales, selling, etc.) | 6927 (25.6%) | 3068 (22.2%) | 3859 (29.2%) |
| Mainly labor (work at production sites, physical work, nursing care, etc.) | 6641 (24.6%) | 3850 (27.9%) | 2791 (21.1%) |
| Current smoker, % | 7004 (25.9%) | 4855 (35.1%) | 2149 (16.3%) |
| Do you telecommute? (Almost never) | 21276 (78.7%) | 10453 (75.7%) | 10823 (81.9%) |
| Have you resigned or changed jobs since April 2020? (Yes) | 1721 (6.4%) | 803 (5.8%) | 918 (6.9%) |
| Do you need any consideration or support from your company to continue working in your current health condition? (Yes) | 6775 (25.1%) | 3338 (24.1%) | 3437 (26.0%) |
| Have you been infected with COVID-19? (Yes) | 195 (0.7%) | 101 (0.7%) | 94 (0.7%) |
| WFun≥21 | 5791 (21.4%) | 2786 (20.2%) | 3005 (22.7%) |
| K6≥5 | 10817 (40.0%) | 4779 (34.6%) | 6038 (45.7%) |
| K6≥10 | 5049 (18.7%) | 2249 (16.3%) | 2800 (21.2%) |
| K6≥13 | 2460 (9.1%) | 1055 (7.6%) | 1405 (10.6%) |
| Perceived poor self-rated health | 13580 (50.2%) | 6732 (48.7%) | 6848 (51.8%) |

## Discussion

We conducted an Internet-based health survey in workers during the third wave of COVID-19 infection in Japan in December 2020. Workers were asked about their socioeconomic status, health status, work status, infection prevention behaviors, and socio-psychological factors.

Internet surveys have become more common in recent years in the fields of public health and epidemiology because relatively large amounts of data can be collected in a short period of time. Compared with conventional population- and workplace-based surveys, Internet surveys have several advantages: it is easier to achieve the target sample size, it is possible to incorporate a large number of batteries, and they can be conducted in a short period of time. In this case, an Internet survey was necessary because the aim was to conduct an urgent study during a phase of rapid spread of COVID-19 infection in Japan. We think our data are valuable for studying working conditions and worker health during the spread of infection.

One of the drawbacks of Internet surveys is the issue of fraudulent responses[18,19]. By answering questions, Internet monitors receive an incentive in the form of points, which have monetary value. This can cause some to provide random or fraudulent responses to earn points; thus, it is important to exclude such respondents. In this survey, we used several algorithms to detect fraudulent responses. First, we included a staged question that asks respondents to choose the third largest number from five numbers. A total of 93% of respondents provided the correct answer for this question. Second, the time taken to answer the question was recorded by the system. Third, answers from respondents with extremely low body weight or short height were judged to be incorrect. Because height and weight questions required the respondents to type in numerical values using a keyboard, we assumed that fraudulent responses were more likely to occur in these questions than in simple click-and-answer questions. Fourth, we examined responses for inconsistencies among questions that were repeated throughout the survey. Questions used to verify inconsistencies inquired about the presence or absence of family members living together and the area of residence. Of 33,087 respondents, 27,036 were judged to have responded appropriately. We confirmed that those who were found to have provided fraudulent responses under one of the four conditions above also often provided fraudulent responses under the other three conditions.

In addition, we were able to increase the credibility of the data by confirming already known relationships between factors. For example, men were more likely to smoke than women, and women were more likely to have higher K6 scores. Region 5, the Kanto region, which includes Tokyo, had more high-income earners than the other regions. There was also more telecommuting experience in Region 5 than in the other relatively rural regions. Moreover, 195 (0.7%) of the 27,036 respondents reported that they had been infected with COVID-19. Because of the self-reported nature of the survey, the data should be interpreted with caution; however, the fact that the lowest infection rate of 0.6% was observed in Region 1, while the highest rate of 1% was in Region 5 are consistent with regional infection rates suggest the validity of this data.

The sampling plan was very important in this study. Workers’ work environment, socioeconomic status, and COVID-19 infection status, which comprised the objective variables of this survey, were expected to vary greatly by region and occupation. In contrast, we assumed that most of the pre-registered respondents in the Internet monitor would reside in urban areas, and that most of the respondents would be office workers. Therefore, we sampled respondents such that they were balanced in terms of sex, type of work, and region in which infection was confirmed.

Selection bias is unavoidable in Internet surveys. This is because respondents are not representative of any group[18,19]. In addition, respondents to Internet surveys are thought to be subject to the volunteer effect due to self-selection for participation. Therefore, it is important to determine the characteristics of the target population of this study by comparing a variety of factors with those in previous studies. The present study collected information on lifestyle-related factors such as smoking, alcohol consumption, and exercise and physical activity. In addition, we employed many health and work-related psychosocial batteries in this study, including K6, the Job Content Questionnaire, Utrecht Work Engagement Scale, WFun, self-rated health, and CDC HRQOL4. All of these have been employed in many workplaces and populations in previous studies.

K6, for example, has been used in many studies. K6 was developed by Kessler et al. to screen for psychiatric distress, such as that observed in depression and anxiety, and is widely used in surveys of the general population as an indicator of mental health[20]. Depending on the survey, cutoff values of 5, 10, and 13 points are used for K6. In the 2007 National Survey on Basic Living Conditions, 27% of male and 33% of female workers had a K6 score of 5 or higher[21]. In a survey of multiple workplaces, 10.8% of 1,709 workers had K6 scores of 13 or higher[22]. In the present study, 40%, 19% and 9.1% had a K6 score of 5 or higher, 10 or higher, and 13 or higher, respectively. These results suggest that while more subjects in this study experienced mild to moderate psychological distress than those in previous studies, they did not show extremely poor health.

In conclusion, this study describes the protocol used to conduct an Internet-based health survey in workers and a summary of its results in December 2020, when COVID-19 was spreading rapidly in Japan. In the future, we plan to use this survey to examine the impact of COVID-19 on workers’ work styles and health.

## Data Availability

No data availability

## Acknowledgements

This study was funded by a research grant from the University of Occupational and Environmental Health, Japan; a general incorporated foundation (Anshin Zaidan) for the development of educational materials on mental health measures for managers at small-sized enterprises; Health, Labour and Welfare Sciences Research Grants: Comprehensive Research for Women’s Healthcare (H30-josei-ippan-002) and Research for the establishment of an occupational health system in times of disaster (H30-roudou-ippan-007); and scholarship donations from Chugai Pharmaceutical Co., Ltd.

Present members of the Collaborative Online Research on Novel-coronavirus and Work (CORoNaWork) project are: Dr. Yoshihisa Fujino (current chairperson), Dr. Akira Ogami, Dr. Arisa Harada, Dr. Ayako Hino, Dr. Chimed-Ochir Odgerel, Dr. Hajime Ando, Dr. Hisashi Eguchi, Dr. Kazunori Ikegami, Dr. Keiji Muramatsu, Dr. Koji Mori, Dr. Kyoko Kitagawa, Dr. Masako Nagata, Dr. Mayumi Tsuji, Dr. Rie Tanaka, Dr. Ryutaro Matsugaki, Dr. Seiishiro Tateishi, Dr. Shinya Matsuda, Dr. Tomohiro Ishimaru, Dr. Tomohisa Nagata, Dr. Yosuke Mafune, and Ms. Ning Liu, in alphabetical order. All of the members are affiliated with the University of Occupational and Environmental Health, Japan.

